# Handwashing Practices among New Mothers and their Guardians: A Mixed-Methods Observational Study in Healthcare Facilities and Households in Rural Malawi

**DOI:** 10.1101/2024.10.11.24315284

**Authors:** Kondwani Luwe, Kondwani Chidziwisano, Katherine Davies, Tracy Morse, Robert Dreibelbis

**Affiliations:** Centre for Water, Sanitation, Health and Appropriate Technology Development (WASHTED), Malawi University of Business and Applied Sciences; Department of Civil and Environmental Engineering, University of Strathclyde; Department of Disease Control, London School of Hygiene & Tropical Medicine (LSHTM)

## Abstract

**Background:** Patient guardians perform hygiene-related roles during postnatal care but are often overlooked in hygiene Interventions. This study examined perceived facilitators and barriers to handwashing behaviour among Malawian new mothers and their guardians in healthcare facilities and households.

**Methods:** This cross-sectional study was conducted in Postnatal Care (PNC) wards (n=2) and households (n=20) in two districts in Malawi. In the PNC wards, 15 mother/guardian pairs were observed and interviewed. In the households 20 new mothers were observed and interviewed while 15 of their guardians were interviewed and/or observed. Water, sanitation and hygiene facilities, handwashing opportunities and actions were documented and analysed using Stata. Behaviour determinants were assessed using the Capabilities, Opportunities and Motivation model in NVivo 14.

**Results:** PNC wards had Handwashing Facilities (HWFs) without soap. Hand rinsing with water only was observed in 20% of all hand hygiene opportunities in the wards (n = 41), with guardians practising it more than mothers. 90% of households lacked HWF. Baby care activities were integrated with chores. Hands were rinsed with water only in 38% of all hand hygiene opportunities (n = 128); before eating (91%), preparing food (36%) and breastfeeding (13%).

Participants knew the importance of handwashing but mothers in wards reported being too tired to get out of bed to wash their hands. The HWFs presented opportunities for handwashing but practice was limited by intermittent water supply, restricted access and soap absence. Participants expressed motivation to buy soap but didn’t prioritise it for handwashing. Mothers often prioritised pacifying their crying newborns and forgot to wash their hands. Guardians were frequently excluded from health promotion activities at the facility.

**Discussion:** Context-specific behaviour change interventions among new mothers and their guardians are needed. Utilising guardians’ support; placing HWFs and reminders strategically; and having innovative approaches to promote soap use should be promoted.

## 1.0 Introduction

Every year across the globe, approximately 2.4 million neonatal deaths occur during the first month of life and 287,000 maternal deaths occur during and following pregnancy or childbirth (1,2). Sub-Saharan Africa suffers a disproportionate burden of these deaths; over half of all neonatal and maternal deaths occur in the region(3). Infection remains a leading cause of these deaths contributing to approximately fifty and ten percent of global infant and maternal deaths respectively (4,5). World Health Organization (WHO) guidelines recommend handwashing with soap at critical times specific to neonatal care: before and or after touching newborns or mothers, caring for the cord, preparing food, breastfeeding, changing the diaper and using the toilet (6–11).

Interventions aimed at improving handwashing with soap during the postnatal period often focus on health workers (12,13) and mothers (14,15) within healthcare facilities. Postnatal care however extends to the household (16–21). Patient guardians are important participants in this continuum of care (14,15,22). Patient guardians - usually relatives, neighbours or friends to patients - are an important part of the health system in low- and middle-income countries (LMICs) (23). They provide person-centred care in healthcare facilities and households such as administering drugs, cleaning and dressing wounds, cooking and serving food, doing laundry, and caring for infants (cord care and nappy changing) (24–27). Despite their significance, few infection prevention and control (IPC) guidelines acknowledge their role or include them in IPC strategies, health education programmes or interventions during postnatal care (28).

The few interventions that target new mothers and their guardians in healthcare facilities and households still face challenges relating to handwashing infrastructure/consumables and behaviour. In both healthcare facilities and homes, handwashing facilities (buckets, jugs, basins, sinks and tippy taps) are limited or non-functional and rarely located in a convenient position (next to patient beds, toilets or kitchens) (20,27,29–33). Handwashing practice also remains low (15,20,34,35) and is influenced by a range of behavioural determinants including knowledge of handwashing with soap; risk perception of disease; protection of infants (nurture); social-cultural norms or perceived social approval (affiliation); forgetfulness; time constraints; and lack of soap due to financial constraints (15,36–42).

Studies that further document handwashing behaviours and their determinants during the postnatal period are still warranted; especially those that extend to the household setting and incorporate the patient’s guardians. This observational study examines handwashing practices and their determinants among Malawian new mothers and their guardians in healthcare facilities and home environments. The results can inform interventions and IPC strategies that are more effective for their similar local contexts.

## 2.0 Methods

### 2.1. Study setting

The study was conducted in Malawi, a country in Sub-Saharan Africa with a neonatal mortality rate of 19 deaths per 1,000 live births and a maternal mortality ratio of 381 deaths per 100,000 live births (43,44). The research took place in Postnatal Care (PNC) wards and households in the catchment areas of two healthcare facilities in the Southern region of Malawi, one being managed by the government in Mwanza district and the other by a Faith-Based Organisation (FBO) in Phalombe district. Both were randomly selected from a list of major healthcare facilities, and each had a capacity of 32 beds in the PNC wards. However, the government healthcare facility was reported to handle an average of more deliveries per week (79 normal deliveries) compared to the one operated by the faith-based organisation (28 normal deliveries). Data were collected at both the healthcare facilities (PNC wards) and households to comprehensively understand handwashing practices and factors that influence the continuum of care for both new mothers and their guardians (45,46).

### 2.2. Study design

This mixed-method observational study followed an explanatory sequential design in which quantitative observations (direct and structured) informed subsequent in-depth qualitative data collection (semi-structured interviews) at the healthcare facility and household.

### 2.3. Study population and sampling

New mothers and their guardians in the PNC wards were conveniently sampled and approached to participate in the study for observations and interviews. At the household, new mothers who had given birth in less than 28 days (different from those targeted in the PNC wards) were randomly selected from the birth register, followed up (with assistance from health staff) and approached to take part in the study for both observations and interviews. Their guardians who had helped care for them while at the PNC ward were also recruited. The study included mothers who underwent a normal delivery and were at least 18 years of age at the time of delivery.

### 2.4. Data collection

Data collection occurred between November 2022 and January 2023 through quantitative methods (checklists and structured observations) and qualitative methods (semi-structured interviews).

#### 2.4.1. Quantitative data collection

##### Facility checklists

Facility checklists focusing on water, sanitation, and hygiene infrastructure and materials were collected at both facilities and at the households of all mothers who participated in household data collection (n=20). The checklists assessed water accessibility (availability of water on or off the premises, distance to water source and the type of water source), toilet facilities (presence, type of toilets and their cleanliness), and handwashing facilities (HWFs) (presence/ absence, type of facilities and availability of handwashing materials such as soap).

##### Structured observations

Observations were conducted with new mothers and their guardians in PNC wards at both facilities. The observations, which lasted three hours (9am to 12 noon), were conducted by two female enumerators. One enumerator was assigned to observe the new mother and the other to observe the guardian. Hand hygiene opportunities were recorded along with associated hand hygiene behaviours – specifically if soap was used; if only water was used; or if hands were not washed at all. The type of handwashing facility used was also recorded together with other relevant notes about the observed practice e.g. the context and activities done before and after the observed critical times for handwashing.

Similar observations were conducted in households, primarily on new mothers and lasted for approximately 4 hours (9 am to 1 pm). In cases where guardians resided in the homes of the targeted new mother, they were also observed.

#### 2.4.2. Qualitative data collection

##### Semi-structured interviews

After the observations, the female enumerators who conducted the observations met with other enumerators (male) and a member of the study management team (KL) to debrief on the findings. Immediately after the debrief, semi-structured interviews were conducted with mothers and guardians. Interviews were conducted in the local language (Chichewa) by the two male enumerators with Bachelor’s degrees in Environmental Health, and prior experience in conducting qualitative research. They all received a week-long training specific to the current study. At the healthcare facility, the interviews were conducted either within or outside the wards, depending on the situation concerning privacy. The participants were asked about their handwashing practices such as barriers and enablers, and reasons for or against washing their hands.

### 2.5. Data processing and analysis

The data collected from checklists and observations using Kobo collect was analysed using StataBE 17 (Stata Corp, College Station, TX, USA). Descriptive statistics were used to obtain the means and frequencies or proportions of continuous and discrete data respectively.

The interviews were recorded, transcribed, and translated into English. Determinants of handwashing behaviours were assessed using the Capabilities, Opportunities, and Motivation model of Behaviour change (COM-B) (47) in NVivo 14(QSR International, Lumivero). The research team organized the responses from the participants into the main areas of the COM-B framework. These were then reviewed and categorized as either facilitators or barriers for postnatal care and household settings. Key themes from the results were identified and combined for both settings. In addition, some field notes from observations were used to further explain and validate the overall results.

### 2.6. Ethical considerations

Ethical approval for the study was obtained from the London School of Hygiene and Tropical Medicine, and the National Committee on Research in the Social Sciences and Humanities (Malawi) (NCST/RTT/2/6., P.09/22/672). Before collecting any data, each participant was asked to provide voluntary written consent. To ensure privacy, female enumerators were assigned to perform observations in the PNC ward, as some women had recently given birth.

## 3.0 Results

### 3.1. Demographics

A total of 35 mothers and 30 guardians participated in the study. This included PNC ward observations and interviews from 9 mother/guardian pairs from the government-run healthcare facility and 6 from the FBO-operated healthcare facility. For household observations and interviews, 10 mothers participated from the government-run facility in Mwanza district and 10 from the FBO-operated healthcare facility in Phalombe district. In 8 of the 20 households, the guardian at the time of study lived with the mother and the guardian participated in both the observations and the interviews. In 7 of the 20 households, the guardian resided elsewhere and was only interviewed. In the remaining 5 of the 20 households, the guardians were not available to participate.

The mothers were generally younger (mean age 24.4 years, SD = 7.2) than the guardians (mean age 42.5 years, SD 10.3). All new mothers had at least completed primary education; 33.3% of guardians had no formal education. Most of the new mothers (97%) were married, while approximately a third of the guardians were widowed or divorced (34%) (Table 1).

**Table 1:** Demographics for the New Mothers and Guardians across both healthcare facilities.

| Demographic |  | PNC (n =30) |  | Household (n =35) |  |
| --- | --- | --- | --- | --- | --- |
|  |  | Mother (n =15) | Guardian (n =15) | Mother (n =20) | Guardian (n =15) |
| Age | Mean (SD) | 23.2 (6.2) | 44.2(10.9) | 25.6(8.1) | 40.9(9.2) |
| Education % | No education | . | 46.7 | . | 20.0 |
|  | Primary | 73.3 | 46.7 | 50.0 | 46.7 |
|  | Secondary | 26.7 | 6.7 | 50.0 | 33.3 |
| Marital status % | Single | 6.7 | . | . | . |
|  | Married | 93.3 | 53.3 | 100 | 66.7 |
|  | Widow | . | 20.0 | . | 13.3 |
|  | Divorced | . | 26.7 | . | 20.0 |

### 3.2. WASH conditions and hand hygiene in PNC wards

#### 3.2.1. WASH conditions at the PNC wards

Both facilities had piped water supply into the PNC ward. Each PNC ward had three pour flush toilets but were generally cleaner in the FBO than in the government-operated health facility. The PNC ward toilets in the government-operated health facility had piped water to sinks with drains for handwashing facilities however there was no running water at the time of the study. In the FBO-run facility, a standpipe outside the toilets was used as a handwashing facility and had running water at the time (Table 2).

**Table 2:** WASH facilities at the PNC wards across the two sites.

| WASH facilities at the PNC |  | Government-run<br>Healthcare facility | FBO-run<br>Healthcare facility |
| --- | --- | --- | --- |
| **Location: Toilets within the PNC wards** |  |  |  |
| Number of toilets |  | 3 | 3 |
| Type of Toilets | Pour Flush | 3 | 3 |
| Condition of toilets | Clean | 2 | 3 |
|  | Unclean | 1 | . |
| Number of handwashing facilities |  | 3 | 3 |
| Handwashing facilities type | Sink with drain | 3 | . |
|  | Standpipe | . | 1 |
| Hand washing materials at HWFs | Water only | . | 1 |
|  | No water | 3 | . |
|  | Soap | . | . |
| **Location: PNC ward accommodation or sleeping area** |  |  |  |
| Number of handwashing facilities |  | 7 | 2 |
| Handwashing facilities type | Sink with drain | 6 | 2 |
|  | Bucket with tap | 1 | . |
| Handwashing materials at handwashing facilities | Water only | 5 | 2 |
|  | No water | 2 | . |
|  | Soap | . | . |
| Posters on handwashing | Present | 5 | 2 |
|  | Absent | 2 | . |

The government-run facility had 7 handwashing facilities (6 sinks and 1 bucket with tap) within the PNC ward and the FBO-operated facility had 2 (sinks); in both facilities, soap was not available. Both PNC wards had different posters at HWF stations and the general ward area, which displayed messages on steps, critical times, and general reminders for handwashing with soap. (Table 2).

#### 3.2.2. Hand hygiene opportunities and actions in PNC wards

Although handwashing facilities were available in the PNC wards of both healthcare facilities, very few people utilised them. A total of 41 handwashing opportunities were observed across all observations (Table 3). Few handwashing opportunities were recorded because most mothers were inactive and spent most of their time sleeping with their guardians sitting beside them. Occasionally, the mothers would wake up to breastfeed their babies while sitting on their beds, usually in response to the baby crying and being handed over to them by the guardians, or after a nappy change. Guardians would also help change baby nappies and dispose of them in a basin or bin.

**Table 3:**
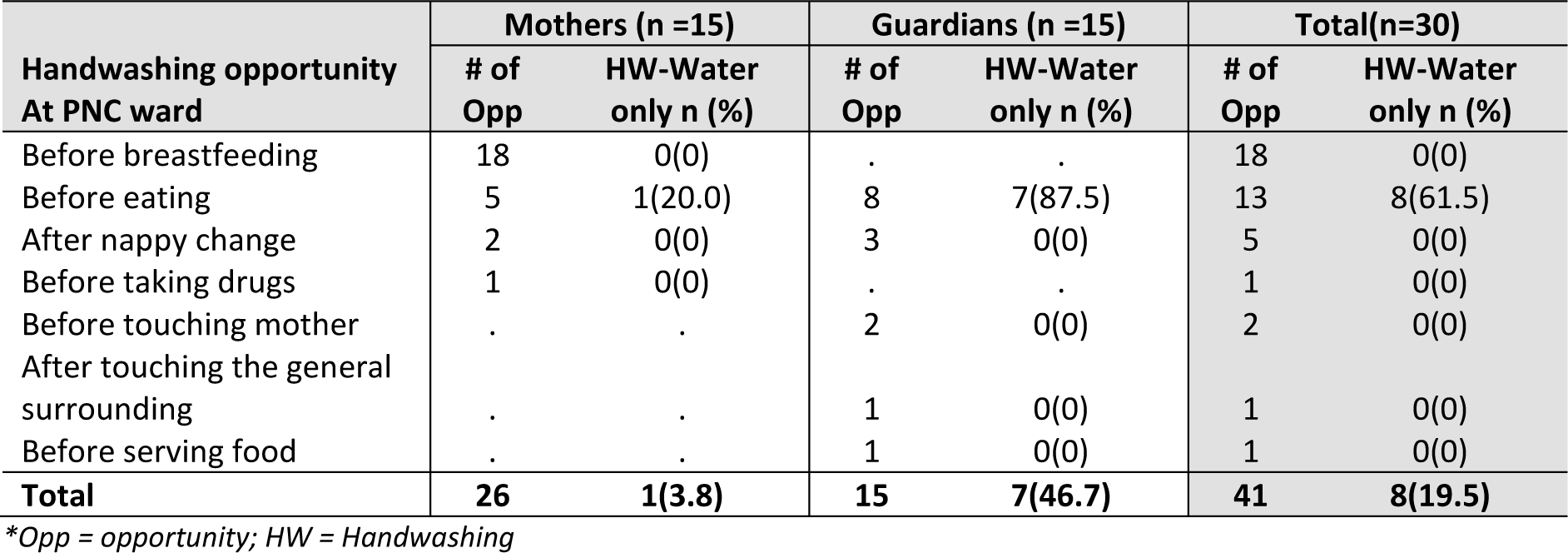
Observed Handwashing opportunities and actions taken by mothers and their guardians at the healthcare facilities.

As soap was unavailable, mothers and guardians washed their hands with water only (hand-rinsing). Hand rinsing was observed in 8 (20%) of all observed handwashing opportunities in the PNC ward and was only practiced before eating. Guardian*s* rinsed hands more often (47%; 7/15) than mothers (4%; 1/26). No mothers washed their hands before breastfeeding or after changing a nappy. The participants used “unconventional” handwashing facilities, such as bottles, jugs and basins (in 6 instances), despite the presence of dedicated handwashing facilities (sinks) within the sleeping areas of the wards in both districts. Available HWFs in the PNC ward were only used in 2 of the 8 observed hand rinsing events. In instances where bottles were used, water was poured into plates or the ground if it was outside the PNC ward.

### 3.3. WASH conditions and hand hygiene in households

#### 3.3.1. WASH conditions at the households

Fourteen out of 20 households across both districts collected their water off-premises, mainly from boreholes which were located within a 500m radius. The majority of the households in Mwanza (n=6) and Phalombe districts (n=6) had traditional pit latrines without slabs. There were dedicated handwashing facilities in only two households which had piped water into their yard, both in Mwanza district. Only one household had water and soap (liquid) at their handwashing facility (Table 4).

**Table 4:**
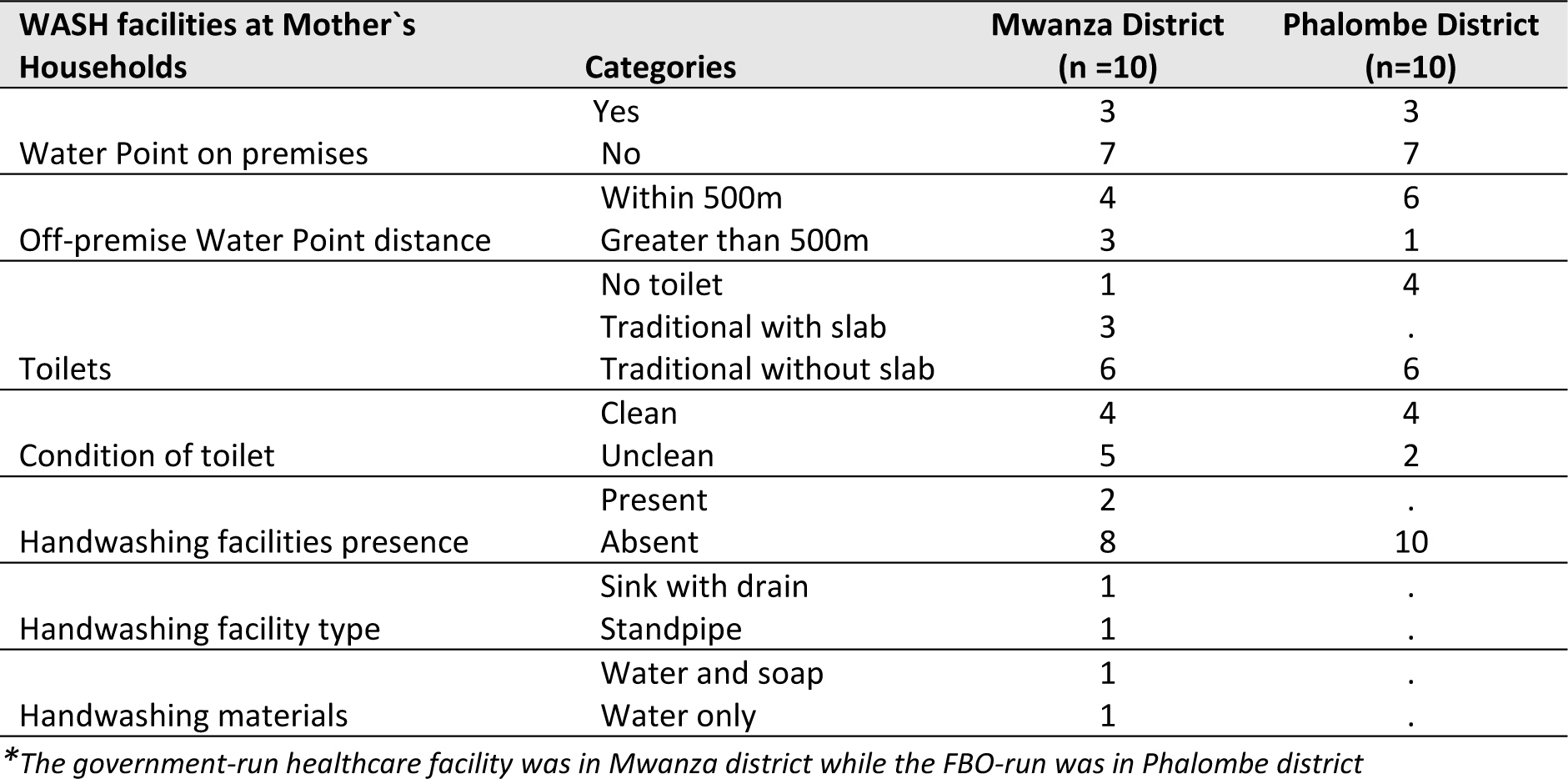
WASH conditions at new mothers’ households across the two study sites.

#### 3.3.2. Hand hygiene opportunities and actions in households

A total of 148 hand hygiene opportunities were observed in households. There was no observed handwashing with soap during structured observations. Hand rinsing practice was done in 49 (38%) of the opportunities, mainly before eating (92%; 23/25) and food preparation (38%; 14/37). Hand rinsing during food preparation was mostly done after making a fire using charcoal resulting in visibly dirty hands. Hand rinsing practice was also observed before breastfeeding (13%; 6/48) and after nappy changes (21%; 3/14) (Table 5).

**Table 5:** Observed Handwashing opportunities and actions taken by mothers and their guardians at the healthcare facility and households.

| Handwashing opportunity Household | Mother (n = 20) |  | Guardian(n=8) |  | Total (n =28) |  |
| --- | --- | --- | --- | --- | --- | --- |
|  | # of Opp | HW-Water only n (%) | # of Opp | HW-Water only n (%) | # of Opp | HW-Water only n (%) |
| Before breastfeeding | 48 | 6(12.5) | . | . | 48 | 6(12.5) |
| Before eating | 23 | 21(91.3) | 2 | 2(100) | 25 | 23(92.0) |
| Before food preparation | 25 | 9(36.0) | 12 | 5(41.6) | 37 | 14(37.8) |
| After nappy change | 14 | 3(21.4) | . | . | 14 | 3(21.4) |
| Before cord care | . | . | 1 | 0(0) | 1 | 0(0) |
| After toilet use | 1 | 1(100) | . | . | 1 | 1(100) |
| After household chores | 2 | 2(100) | . | . | 2 | 2(100) |
| **Total** | **113** | **42(37.1)** | **15** | **7(46.7)** | **128** | **49(38.3)** |

\*Opp = opportunity; HW = Handwashing

**Table 6:** COM-B facilitators and barriers to handwashing with soap among new mothers and guardians in healthcare facilities (PNC) and households (HH).

| COM-B Components | Facilitators | Barriers |
| --- | --- | --- |
| **Physical Capability** | Some messages and skills from PNC health talks (PNC) | Irregular PNC health talks (PNC)<br>Tiredness or feeling pain after giving birth (PNC) |
| **Psychological Capability** | Knowledge of handwashing with soap and disease prevention (PNC and HH)<br>Knowledge of handwashing with soap at critical times (PNC and HH) |  |
| **Physical Opportunity** | Presence of Hand washing facilities (PNC)<br>Water availability (PNC and HH) | Handwashing facility absence and related issues (HH and PNC)<br>Intermittent water supply (PNC and HH)<br>Soap access and affordability (PNC and HH) |
| **Social Opportunity** | Guardians' support in handwashing or Guardians bringing handwashing facilities to Mothers (PNC and HH)<br>Health staff support in handwashing (PNC)<br>Income source and decision-making by self (PNC and HH) | Income source and decision-making by others (PNC and HH)<br>Absence of Guardians during health talks (PNC) |
| **Automatic Motivation** | Clean sanitary facilities (PNC)<br>Handwashing to prevent diseases (PNC and HH) | Disgust due to unhygienic sanitary facilities (PNC)<br>Forgetfulness, hunger, negligence for handwashing with soap (PNC and HH)<br>Prioritisation of nurturing pacifying baby over handwashing (PNC and HH) |
| **Reflective Motivation** | Money use - prioritisation for the purchase of soap (PNC and HH)<br>Money use - prioritization for the purchase of buckets and handwashing items (PNC and HH) | No soap use prioritisation for handwashing (PNC and HH)<br>No soap was provided by the hospital (PNC)<br>The perception that using sinks for handwashing is inappropriate (PNC) |

\*PNC denotes issues mentioned at the healthcare facility Level and HH denotes issues mentioned at the household level.

The use of “unconventional” handwashing facilities was also observed at the household level. Compounded by a lack of dedicated handwashing facilities, the participants used the following items for hand rinsing in the 49 recorded instances: basins (51%); Jug and Basin (14%); cups (14%) and buckets without tap (10%) respectively. Taps, pots and plates were used in fewer instances. Participants dipped their hands in water when using basins, buckets without taps, pots and plates. In 3 instances multiple users were observed dipping their hands in the same water. Pouring was done when using a jug and basin and cups. Baby care activities (e.g., breastfeeding) at the household level were often integrated with other daily activities like laundry, washing dishes, fetching water and preparing food. Other members of the household (guardians) also helped with other baby care activities such as nappy changing and cord care using an antiseptic (spirit).

### 3.4. Reported determinants of hand hygiene practices

Thematic analysis of the qualitative data found similar findings of determinants of hand hygiene in both PNC wards and the home environment (HH); qualitative results are combined across both settings. We note any setting-specific information in the results.

#### 3.4.1. Capabilities

##### Facilitators

Across most of the interviews, participants demonstrated an understanding of the critical times for handwashing, demonstrating high levels of action knowledge. They were generally aware of specific moments, such as after changing diapers and using the toilet, emphasizing the importance of handwashing.

The participants understood the link between handwashing with soap and disease prevention. They recognised that handwashing can eliminate disease-causing germs. A guardian at the household emphasised this connection, stating:

> *“We normally use (wash hands) soap because we want to get rid of the germs that can be in our hands from baby’s diapers”* [Participant 8, Guardian, Mwanza, Household].

Information about when to wash hands was also repeated during health talks received at the facility, one mother at the PNC ward said:

> *“They [Health staff] encouraged mothers to wash hands before breastfeeding the baby, and after changing a diaper”* [Participant 6, Mother, Phalombe PNC ward].

##### Barriers

Participants reported that the health talks were not regular and mainly done on discharge, as such some reported not getting hand hygiene messages during their stay in the PNC wards.

The reported fatigue and pain experienced by new mothers in the PNC ward created additional barriers. The physical strain made it challenging for them to focus on messages (when given out) and practice handwashing. One participant said:

> *“The guardian brought me spaghetti for lunch but I ate without washing my hands because I was too lazy to wake up and walk to the sink to wash my hands, so I just started eating with unwashed hands”* [Participant 5, Mother, Phalombe, PNC ward]

#### 3.4.2. Opportunities

##### Facilitators

In terms of physical opportunity, sinks with running water were available in all PNC wards but participants did not use them partly due to restrictions. They however showcased “resourcefulness” in utilising other means for handwashing (such as bottles and basins) at PNC wards.

The use of bottles and basins was also eminent in the households. One guardian in explaining how they washed hands at the household said:

> *“Mostly using a bottle, we ask someone to help pour water over hands and wash hands.”* [Participant 6, Guardian, Phalombe, Household].

This habit from the household environment could also explain the shunning of using the sinks at the PNC wards

Guardians were also an important component of social opportunity for handwashing with soap – both at the facility and at home; supporting mothers with HWFs and assisting in handwashing practices. A guardian at the PNC ward confirmed their role, stating:

> *“My main role here is to help the mother take care of the baby, such as holding the baby while the mother is taking a bath, giving her water to wash her hands and breasts before carrying the baby”* [Participant 9, Guardian, Mwanza, PNC ward].

The observation of health workers practising proper hand hygiene further served as a social opportunity for mothers and their guardians to adopt similar practices. Participants reported witnessing health staff adhering to hand hygiene practices, such as glove use in the delivery room and PNC ward.

##### Barriers

Even though guardians were an important component of the social opportunity, they were separated from the new mothers and were absent during health talks. The absence of guardians during health talks limited information dissemination and potentially hindered the reinforcement of handwashing practices by the guardians. This separation was reported to be justified by health workers as a means to avoid disturbance during work hours. A guardian at the PNC wards reported:

> *“Guardians are not given any information; they are sent out when the mothers are given such information at the time of discharge”* [Participant 3, Guardian, Mwanza, PNC ward].

Participants also reported that few sinks were functional and would at times be restricted from using them. A mother at the PNC ward said:

> *“We were not allowed to use them because they said some people were not using them properly, for example, washing plates which make the sinks broken with the food scraps”* [Participant 1, Mother, Mwanza, PNC ward].

This possibly added further to the observed shunning of using the sinks for handwashing.

Intermittent water supply both at the PNC ward and household levels, emerged as an obstacle. This was reported to be intense during the dry season in the households. Participants also raised concerns about the intermittent water supply at the PNC wards, with one stating:

> *“Water supply is not continuous; the water doesn’t take long before it stops.”* [Participant 3, Guardian, Phalombe, PNC ward]

Much as some participants would afford to buy soap, its high cost and unavailability at handwashing facilities in the wards acted as a barrier to consistent handwashing practice. Some cited financial constraints as a reason for not being able to buy soap, as expressed by one guardian at the household:

> *“To tell the truth, soap is hard to find because of the small amount I earn; the priority is to buy food to feed my children”* [Participant 5, Guardian, Phalombe, Household].

#### 3.4.3. Motivation

##### Facilitators

Several motivational factors were identified that had the potential to encourage handwashing practices. Some reported that they washed their hands to prevent diseases; this emanated from their facilitative psychological capability of knowing the link between handwashing with soap and disease prevention.

Participants prioritised money for the purchase of handwashing-related items (soap and buckets). One guardian at the PNC ward said:

> *“Yeah, I buy household utensils such as plates, basins, buckets, and everything you know a woman needs in the household”* [Participant 6, Guardian, Phalombe, PNC ward].

The participants’ reports of allocating financial resources for such purchases, even while at the PNC ward, showed their reflective motivation to maintain hygiene during the postnatal period. Participants also considered soap as a priority purchase even in households, as expressed by one Mother:

> *“So, the money we find we buy food such as maize, we buy soap, clothes for children, paying school fees”* [Participant 4, Mother, Mwanza, Household].

This is however negated later; the soap was not prioritised for handwashing as outlined in the barriers, and handwashing with soap was not observed in the study.

The cleanliness of PNC wards, especially in the FBO-run healthcare facility, also served as a motivational factor. A respondent highlighted this cleanliness, stating:

> *“The hygiene conditions are good because the toilets are very clean, the postnatal care room is very clean that we can eat our foods there without any problem”* [Participant 1, Guardian, Phalombe, PNC ward].

It is important to note that the FBO-run healthcare facility in Phalombe district is a private hospital, unlike the one in Mwanza district which is a government-run hospital and is public.

##### Barriers

Despite these facilitators, barriers to motivation were apparent. Some participants reported discontentment with the cleanliness of the sanitary facilities in the government-run healthcare facility and would not use them. Particularly, the handwashing facilities in the toilets in the PNC had no running water. A mother at the PNC ward said:

> *“Sometimes I am not willing to go there to take a bath because the bathrooms are dirty”* [Participant 7, Mother, Mwanza, PNC ward].

The participants reported that the sanitary facilities were particularly dirty at night even after being cleaned twice a day (morning and evening) - the facilities were not cleaned during the night.

Some people avoided using HWFs in the PNC ward due to the perception that sinks ought not to be used for handwashing. For instance, some participants mentioned that they would collect water in a cup from the sink and wash their hands outside the wards to avoid “dirtying the ward/sink.” Others believed it was inappropriate to wash hands in the sinks because they were also used as drinking water collection points. Confirming this, one mother said:

> *“People were washing hands in the sinks but to me, I think it’s not right to wash hands there since we also collect drinking water from the same sinks”* [Participant 4, Mother, Mwanza, PNC ward].

Even though some participants would afford to buy soap, they expressed a challenge in prioritising it for handwashing, opting to allocate this limited resource for other household purposes. One mother at the household stated:

> *“Whenever soap is available, it is used only for washing utensils and bathing, but sometimes where we have no soap, we just take a bath with water only”* [Participant 3, New Mother, Mwanza Household].

A guardian in the PNC ward added on to say:

> *“We try to save it because if we are to use it for handwashing frequently it means it will not last long because we use the same for bathing the patient and washing her clothes”* [Participant 5, Guardian, Phalombe, PNC ward].

The participants also reported instances where they had forgotten to wash their hands, such as after visiting the market, using the toilet, before eating particular snacks, or before holding the baby especially when it was crying. One Mother at the household reflected on her actions and stated:

> *“Like just now, I have just picked up the baby and started breastfeeding and yet at the hospital, they said we have to wash hands before breastfeeding the baby”* [Participant 10, New Mother, Mwanza Household].

Another mother at the PNC ward added to this and said:

> ***“…****if it [the baby] suddenly wakes up and starts crying, I may not think of going to wash hands first but I can just start breastfeeding the baby….”.* [Participant 2, New Mother, Phalombe, PNC ward]

## 4.0 Discussion

The study revealed low compliance with handwashing practices among new mothers and guardians in healthcare facilities and household settings despite generally high capability (knowledge) about handwashing with soap. Opportunity for handwashing emerged as a major barrier to improved hand hygiene – both in terms of low physical opportunity due to limited soap and irregular water and social opportunity due to the exclusion of guardians from health talks during patient discharge. Motivation for handwashing was mixed, with mothers prioritising money for handwashing-related items such as buckets, basins and soap however, soap was not prioritised for handwashing. Mothers also forgot to wash hands when tending to their baby when it was crying. These findings are consistent with existing literature on hand hygiene during the neonatal period, highlighting the challenge of translating knowledge into practice (36,48). The observed missed opportunities for handwashing in both settings showcase the need for targeted interventions to improve handwashing practices (27,35).

The findings also provide insights into the handwashing practices among new mothers and their guardians in healthcare facilities and household settings. Generally, participants utilised fewer opportunities to wash their hands at the healthcare facility than at the household, raising important considerations regarding the environmental and contextual factors influencing hand hygiene behaviours, also raised by Tamene *et al* (2023)(33) and Dreibelbis *et al* (2013)(49).

Participants at the healthcare facility faced limited opportunities for hand washing by being told not to use the handwashing facilities, similar to findings by Mangochi *et al* (2022) in their study on hand hygiene practice in Malawian neonatal healthcare units (50). Healthcare workers are the ones that normally use the HWFs within the PNC wards however this practice limits effective Infection Prevention and control which warrants the involvement of everyone within the healthcare facility including the new mothers and their guardians (51). The limited use of HWFs was also compounded by intermittent water supply and few functional HWFs among others, a common scenario in Low- and Middle-Income countries (50,52–54). Despite being motivated to use hygienic facilities, some participants were hesitant to use the sinks for handwashing because they also served as points for collecting drinking water. They wanted to keep these points clean for drinking purposes. It’s important to recognize that patients in healthcare facilities have limited control over their environment. Therefore, facilities need to ensure that they provide appropriate handwashing facilities that are specific to the context and enable the behaviours they are promoting (51,55).

A refined approach to postnatal care, recognising the physical vulnerability and specific needs of mothers immediately after childbirth is needed (27,30). Mothers were too tired or sore to move from the bed to handwashing facilities that were available in the PNC ward. The “unconventional” HWFs and their use by guardians represent an adaption to this reality, with guardians bringing portable objects to mothers to use to clean their hands. Guardians, therefore, play an important role in facilitating hand hygiene for new mothers (27,37,50,56). However, the separation of guardians from new mothers during crucial health talks indicates a missed opportunity for reinforcing these essential hygiene practices. While most interventions traditionally target healthcare workers and mothers, this finding supports the need for inclusive interventions involving patient guardians or caretakers (12,14,15). Moreover, guardians who are actively involved in various aspects of care can serve as key influencers in promoting handwashing practices (23). Recognizing and reinforcing the role of guardians in the continuum of care is therefore essential (46,57).

The findings regarding handwashing practices within the household further shed light on some key contextual considerations. Mothers displayed varied levels of handwashing, with some engaging in this practice only when they perceived their hands as visibly dirty, especially during cooking activities, as also reported in other studies (37,58). This observation shows a potential gap in awareness and habitual handwashing behaviours within the household setting. Effective handwashing practice is also challenging given the concurrent management of baby care and other household tasks by new mothers. This is a common scenario in the African context and often results in instances where handwashing is overlooked(37,59,60). To promote consistent handwashing practices, interventions could be tailored to these specific dynamics-placing HWFs near areas where caregiving and food-related activities occur could serve as a practical solution (61–65). Implementing reminders, such as visual cues associated with the baby’s care routine, may prompt mothers and guardians to wash their hands regularly (41,60). Other innovative approaches, such as custom-made hand sanitisers attached to baby accessories or worn by mothers, could provide a convenient means for incorporating hand hygiene into the routine of baby care (66). This could prove effective in instances where immediate responses to a baby’s needs take precedence over handwashing.

The identified barrier of limited affordability and prioritisation of soap for other uses other than handwashing should be addressed. In resource-constrained settings, where households may face economic challenges, promoting hand hygiene becomes linked with addressing broader socioeconomic factors (37,48,67). One potential solution for encouraging the prioritisation of soap for handwashing is through approaches that emphasize the dual functionality of soap for personal hygiene and broader domestic purposes (68). Pointing out the role of soap in overall health and well-being, not just hand hygiene may change perceptions of soap as a scarce resource. Such messages may also be integrated into existing hospital or community programs, such as maternal and child health initiatives (68). Moreover, community-based initiatives or partnerships with local organizations could explore ways to make soap more accessible, through for example subsidised programs or collaborations with soap manufacturers (69). Empowering communities to produce low-cost, locally-made soaps could be another option (70).

### 4.1. Limitations of the study

One limitation of the study was the few recorded observed opportunities for handwashing because mothers often slept for extended periods in the PNC wards. This restricted the researchers’ ability to capture a comprehensive picture of handwashing practices throughout the day. Even though this is a reflection of what happens during this period, future studies may benefit from employing alternative data collection methods, for example, the use of cameras or adjusting observation schedules to capture a wider range of activities and behaviours related to hand hygiene. Even though the study employed the support of direct observations to counter this, it is possible that some of the observed behaviours were modified in response to being observed (Hawthorne effect). As with any single-site study, the generalisability of the findings may be limited to the specific context and population under investigation. While the study provides insights into handwashing practices among new mothers and guardians in Malawi, caution should be exercised when extrapolating the results to other settings or populations with different sociodemographic characteristics.

## 5.0 Conclusion

Our study points to the need for targeted interventions to improve hand hygiene practices among new mothers and guardians in both healthcare facilities and household environments. Despite adequate knowledge, compliance with handwashing practices remains low due to limited opportunities and mixed motivations. Factors such as limited access to handwashing facilities, intermittent water supply, soap affordability and use, physical strain on new mothers, and concurrent caregiving and household tasks further complicate handwashing with soap practices. Strategies to address these challenges include empowering and utilising patient guardians, strategic placement of handwashing facilities, implementing reminders, and innovative approaches to promote soap use prioritisation for handwashing. Collaborative efforts involving healthcare providers, communities, and policymakers are essential to implement and sustain interventions aimed at promoting handwashing with soap during the postnatal period.

## Data Availability

All data produced in the present study are available upon reasonable request to the authors

## Acknowledgements

We extend our gratitude to all participants who generously shared their time and experiences, without whom this study would not have been possible. We acknowledge the support and cooperation received from the healthcare facilities and communities involved in this research. Special thanks to the research enumerators for their dedicated efforts in data collection and management.

